# Pregnancy-related Heart Failure Readmissions in the US: Analysis of the Nationwide Readmissions Database

**DOI:** 10.64898/2026.02.05.26345711

**Authors:** Emma Barry, Min Jung Kim, Sarah A Goldstein, Anna E. Denoble, Patricia Chavez, Christine Hsueh, Sara Tabtabai

## Abstract

**Background:** Cardiovascular disease (CVD) is the leading cause of pregnancy-related morbidity and mortality in the United States. Several studies have evaluated readmission rates in the general HF population, but in patients with pregnancy-related HF, readmissions have been understudied. This study aims to characterize the 30-day HF readmission patterns in pregnancy-related admissions to identify vulnerable patient populations.

**Methods:** The National Readmission Database from 2016 to 2021 was used to identify women aged 13-49 with an index hospitalization in which HF was coded as either the primary or secondary diagnosis during a pregnancy-related antepartum, delivery, or postpartum admission, identified by diagnosis-related group (DRG) codes and ICD-10 codes. The primary outcome was 30-day all-cause readmission. We performed descriptive and comparative analyses to describe the differences in patient characteristics and readmission patterns between groups.

**Results:** The overall 30-day all-cause readmission rate was 13% when readmissions for delivery were excluded. The readmission rate increased with age, peaking at 15.1% in the 38-49yr age group. Higher readmission rates were also associated with combined (systolic and diastolic) HF (16.1%), systolic HF (14.8%), lower socioeconomic status (15.3%), substance use disorder (17.2%), and alcohol use (18.6%). Patients whose index hospitalization was for delivery had the highest absolute risk of 30-day readmission at 19.3%. Readmissions peaked between days 6 and 8 post discharge, with more than 50% of all readmissions occurring within the first two weeks post-discharge

**Conclusions:** In our study, the highest risk of readmission occurred after an index hospitalization for delivery, and most readmissions occurred in the first 2 weeks post-discharge. Our findings suggest that a post-discharge follow up within 7 days of admission complicated by HF should be extended to patients with pregnancy-related HF and effective readmission reduction strategies must include a better understanding of heart failure phenotypes, and a proactive approach to addressing social risk factors.

## INTRODUCTION

Cardiovascular disease (CVD) is the leading medical cause of pregnancy-related morbidity and mortality in the United States.^1–4^ Recent data suggests that 10.4% of pregnancy-related deaths are attributed to cardiovascular conditions, including cardiomyopathy.^5^ Cardiac conditions during pregnancy can be either congenital, acquired, or pregnancy-associated.^6^ Prior studies have established CVD as an increasing cause of adverse outcomes in pregnancy.^7–9^ A recent study used the National Inpatient Sample from 2010 to 2019 to evaluate birth hospitalizations and found that patients who had CVD had higher maternal mortality, stillbirth, preterm birth, and congenital anomalies. CVD was also associated with an increase of $2,598 per patient during admission for delivery^1^.

HF in particular has been identified as the underlying cause of over 9% of cardiovascular-related in-hospital deaths among hospitalizations related to pregnancy^10^. In expectant mothers with pre-existing heart disease, HF remains the most common major cardiovascular complication in pregnancy, affecting 11% of such pregnancies.^10–13^ Furthermore, women with a diagnosis of HF were more likely to experience adverse maternal outcomes.^10^ Briller *et al* evaluated HF among pregnancy-related hospitalizations and demonstrated that approximately 60% of HF cases occurred postpartum, followed by delivery and antepartum^10^. Few studies have focused on the risk of readmission related to HF in pregnancy,^14,15^ with most focused on the association with hypertensive disorders of pregnancy.^16,17^ This study aims to characterize the 30-day HF readmission patterns in pregnancy-related admissions to identify vulnerable patient populations.

## METHODS

### Data Source

We conducted a retrospective cohort study using the Healthcare Cost and Utilization Project (HCUP) Nationwide Readmissions Database (NRD) for the years 2016 through 2021. The NRD is one of the largest publicly available all-payer inpatient databases representing 49.1% of hospitalizations in the United States. This database includes all hospitalizations with and without repeat admission in a calendar year and it is composed of clinical and nonclinical variables that support readmission without identification.^18^

### Study Population

We identified women aged 13 to 49 years with an index hospitalization in which HF was coded as either a primary or secondary diagnosis. We subsequently identified those with a pregnancy-related admission, defined as an admission in which a pregnancy-related diagnosis-related group (DRG) or International Classification of Diseases, 10^th^ edition (ICD-10) code was recorded. Since the NRD only includes readmissions within the same calendar year, to ensure complete 30-day follow-up within the same calendar year, analyses were restricted to patients hospitalized between January and November.

### Outcome Measures

The primary outcome was 30-day all-cause readmission, defined as any unplanned inpatient admission to an acute care hospital within 30 days of discharge from the index hospitalization. Planned readmissions for elective procedures were excluded according to HCUP recommendations. In order to account for antepartum admissions with a subsequent readmission for delivery, delivery-related readmissions were defined using established MS-DRG codes for delivery, including cesarean and vaginal deliveries (MS-DRG 765-768, MS-DRG 774-775). These admissions were excluded in a sensitivity analysis.

Secondary outcomes included:

1. Etiology of index admission and readmission, categorized into major diagnostic categories (MDCs) and specific diagnosis-related groups (DRGs).
2. Comorbidity burden, assessed using the Elixhauser Comorbidity Index^18^, individual comorbid conditions, and pregnancy-related comorbidity codes.
3. Admission characteristics, including length of stay (LOS), discharge disposition, hospital charges, and severity/mortality risk subclassifications.
4. Timing of readmission, defined as days from discharge to the first readmission.

### Covariates

Covariates included patient characteristics and clinical characteristics, including age, household income quartile, primary payer (Medicare, Medicaid, private insurance, or other), comorbidity burden, pregnancy related comorbidity), and behavioral risk factors (tobacco use, alcohol use, substance use disorder). Mode of hospital admission was also included, defined using the HCUP emergency department (ED) service indicator to distinguish hospitalizations originating from the emergency department versus those resulting from direct admission by a physician.

Comorbidity burden was assessed using the Elixhauser Comorbidity (EC) classification system, which consists of 29 binary comorbidity indicators derived from ICD diagnosis codes and APR-DRG information listed on the index hospitalization record. Overall comorbidity complexity was quantified using the Elixhauser risk score (ECs), calculated as the sum of the 29 individual comorbidities. In-hospital mortality Elixhauser risk scores (MECs) and 30-day readmission Elixhauser risk scores (RECs) were calculated using the weights proposed by Moore et al.

Mortality risk and severity risk were measured using scores ranging from 0 to 4 derived from the All Patient Refined Diagnosis-Related Group (APR-DRG) methodology. These scores incorporate information from primary and secondary discharge diagnoses, patient age, and preexisting medical conditions documented in discharge billing codes. Discharge disposition was categorized as home or self-care, transfer to another healthcare facility (e.g., short-term hospital, skilled nursing facility, or other facility), home healthcare, or discharge against medical advice.

### Statistical Analysis

We applied HCUP-provided discharge weights to generate nationally representative estimates. Categorical variables were compared between patients with and without 30-day readmission using χ² tests, while continuous variables were compared using t-tests or Wilcoxon rank-sum tests as appropriate. Results are presented as proportions (%) or means ± standard deviation (SD).

The etiologies of index hospitalizations and subsequent 30-day readmissions were characterized using Major Diagnostic Categories (MDCs), which classify inpatient admissions according to the principal diagnosis and underlying organ system or disease etiology. The distribution of MDCs for index admissions and readmissions was summarized descriptively and visualized using bar and pie charts. Timing of readmission was analyzed as the proportion of total readmissions occurring on each day post-discharge. All statistical analyses were performed using SAS 9.4 (SAS Institute, Cary, NC). A two-sided p < 0.05 was considered statistically significant.

## RESULTS

### Study Population and Patient Characteristics

A total of 20,695,094 hospitalizations among women aged 13 to 49 years were identified from the HCUP National Readmission Database (NRD) between 2016 and 2021 (Figure 1). Among these, 48,300 hospitalizations were associated with HF. From this group, 21,200 hospitalizations were identified as involving the pregnancy continuum, including antepartum, delivery, and postpartum admissions.

**Figure 1.**
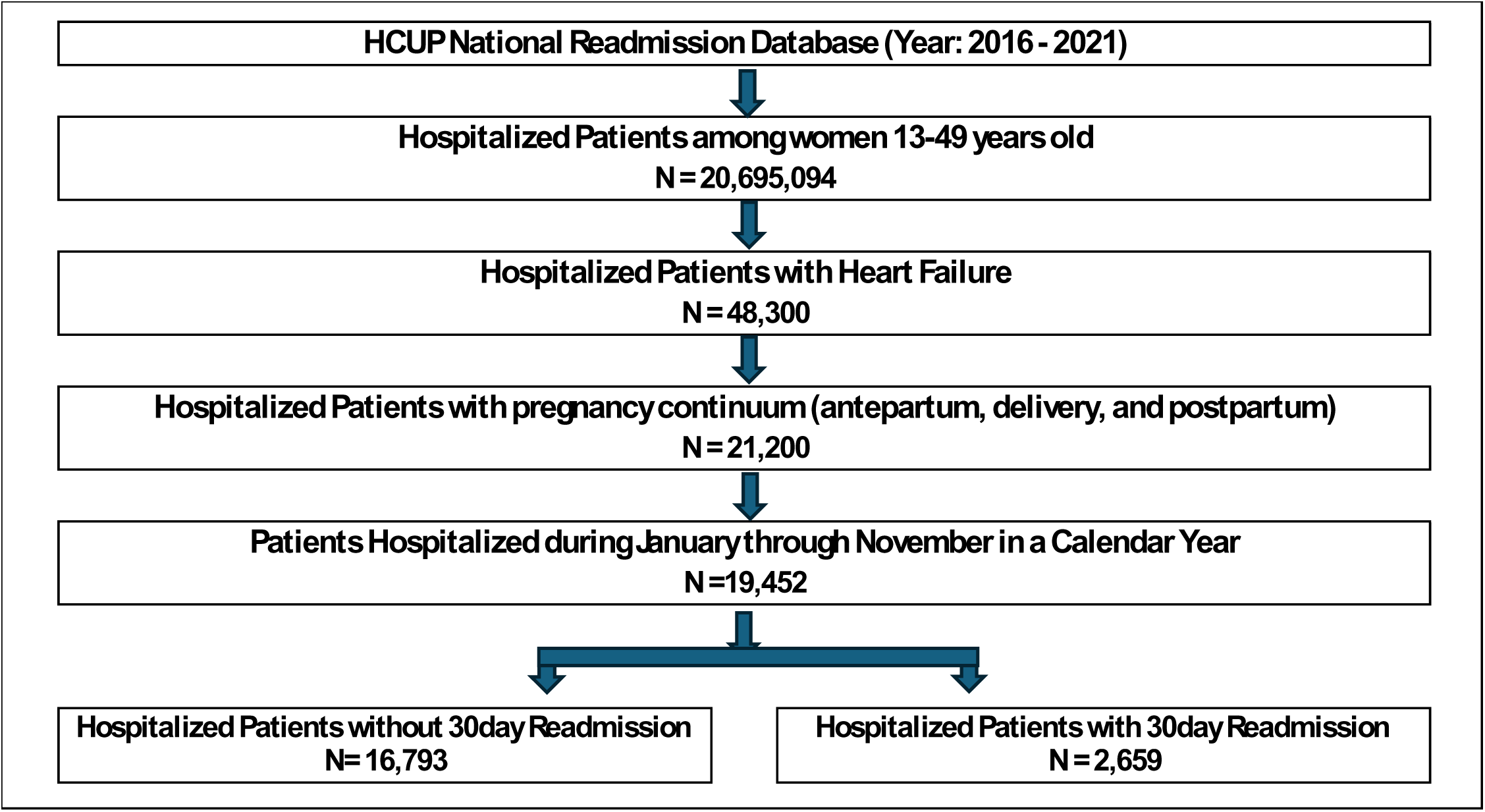
Cohort Selection Flowchart for Pregnancy-Related Hospitalizations With Heart Failure (NRD 2016–2021)

After restricting to hospitalizations occurring between January and November, there were 19,452 eligible hospitalizations. Among these, 2,659 hospitalizations (13.7%) were followed by at least one 30-day readmission.

Patient demographic, clinical, and socioeconomic characteristics stratified by 30-day readmissions are shown in Table 1. The likelihood of HF readmissions increased with increasing age group, with the highest readmissions rates in the 34-37- and 38-49-years cohorts (13.8% and 15.1% respectively, *p=0.005*). Patients in the lower quartile of household income and on Medicare or Medicaid had the highest readmission rates (15.3%, *p=<0.0001*; 23.5%, *p<0.0001*). When stratified by type of heart failure, systolic heart failure and combined heart failure had the highest readmission rates (systolic 14.8%, combined 16.1%, *p=<0.0001)*. Behavioral characteristics were associated with significantly higher readmission rates. Alcohol use resulted in a readmission rate of 18.6%, and substance use 17.2% (*p=<0.0001*), while tobacco use was not associated with a statistically significant difference in readmission.

**Table 1.** Patient Demographic, Clinical, and Socioeconomic Characteristics by 30-Day Readmission Status among Women Aged 13–49 Years with Heart Failure (NRD 2020–2021).

| Patient Characteristics | 30Day Readmission |  |  |  |
| --- | --- | --- | --- | --- |
|  | Without 30Day Readmission | With 30Day Readmission | p_value | Rate (%) |
| Unweighted Index Admission | 16,793 (86.3%) | 2,659 (13.7%) |  |  |
| Weighted Index Admission | 32,768 (86.5%) | 5,102 (13.5%) |  |  |
| Age Group |  |  |  |  |
| 13 - 17 | 0.4% | 0.2% | 0.005 | 7.4% |
| 18 - 28 | 26.7% | 25.0% |  | 12.9% |
| 29 - 33 | 26.9% | 25.5% |  | 13.1% |
| 34 - 37 | 21.5% | **21.7%** |  | 13.8% |
| 38 - 49 | 24.6% | **27.6%** |  | 15.1% |
| HF Subtype |  |  | <.0001 |  |
| Systolic Heart Failure | 41.2% | 45.3% |  | 14.8% |
| *Without PPCM\** | 38.5% | 39.7% | 0.457 | 15.2% |
| *With PPCM\** | 61.5% | 60.3% |  | 14.6% |
| Diastolic HF | 19.8% | 14.9% |  | 10.6% |
| Combined HF | 8.8% | **10.6%** |  | 16.1% |
| Other HF | 30.2% | 29.2% |  | 13.3% |
| PPCM* | 43.45% | 43.14% |  | 13.46% |
| Household Income |  |  |  |  |
| Lower Quartile | 40.5% | **46.1%** | <.0001 | 15.3% |
| Second Quartile | 26.0% | 25.6% |  | 13.5% |
| Third Quartile | 20.9% | 18.5% |  | 12.3% |
| Upper Quartile | 12.6% | 9.9% |  | 11.0% |
| Primary Payer |  |  |  |  |
| Medicare | 8.5% | **16.5%** | <.0001 | 23.5% |
| Medicaid | 54.5% | **57.8%** |  | 14.4% |
| Private (including HMO) | 31.4% | 20.1% |  | 9.2% |
| Others including self-pay | 5.7% | 5.5% |  | 13.4% |
| Patient Location |  |  |  |  |
| Metropolitan ($\geq 1M$) | 54.6% | 54.2% | <.0001 | 13.5% |
| Metropolitan ($< 1M$) | 31.9% | 35.2% | | 14.8% |
| Micropolitan | 7.8% | 6.3% |  | 11.2% |
| Others | 5.7% | 4.3% |  | 10.7% |
| Behavioral Characteristics |  |  |  |  |
| Tobacco Use | 10.1% | 11.1% | 0.12273 | 14.8% |
| Alcohol Use | 6.3% | **9.1%** | <.0001 | 18.6% |
| Substance use disorder | 23.4% | **30.8%** | <.0001 | 17.2% |

*PPCM*\*: Peripartum Cardiomyopathy

When antepartum admissions followed by a delivery admission were excluded, 156 readmissions (5.8% of all readmissions) were found to be delivery-related, corresponding to 0.8% of all index admissions in the cohort. When these delivery-related readmissions were excluded in a sensitivity analysis, the overall 30-day readmission rate decreased modestly from 13.7% to 13.0%, and importantly, the direction and magnitude of associations across key subgroups remained unchanged.

### Peripartum Cardiomyopathy

There was no significant difference in readmissions in patients coded with systolic HF either with or without a designation of PPCM. PPCM alone had no significant difference between readmission and no readmission, and overall readmission rate was 13.46%.

### Index Admission Characteristics

Index admission characteristics stratified by whether a 30-day readmission occurred are shown in Table 2. Index admissions characterized as mortality risk with major likelihood of dying and severity with major loss of function were more likely to result in a readmission, with readmission rates of 16.7% and 15.9%, respectively (p=<0.0001). Index admissions associated with delivery were most likely to be followed by a 30-day readmission compared to other phases of pregnancy, with a readmission rate of 19.3% (*p=<0.0001*). The index hospitalization length of stay (LOS) associated with the highest readmission rate was seen in patients with prolonged index admissions of 6-8 days (readmission rate 15.2%). Regarding discharge disposition, the highest rates of readmission were seen in patients discharged to home health care (17.3%) or discharged against medical advice (33%, *p=<0.0001*).

**Table 2.**
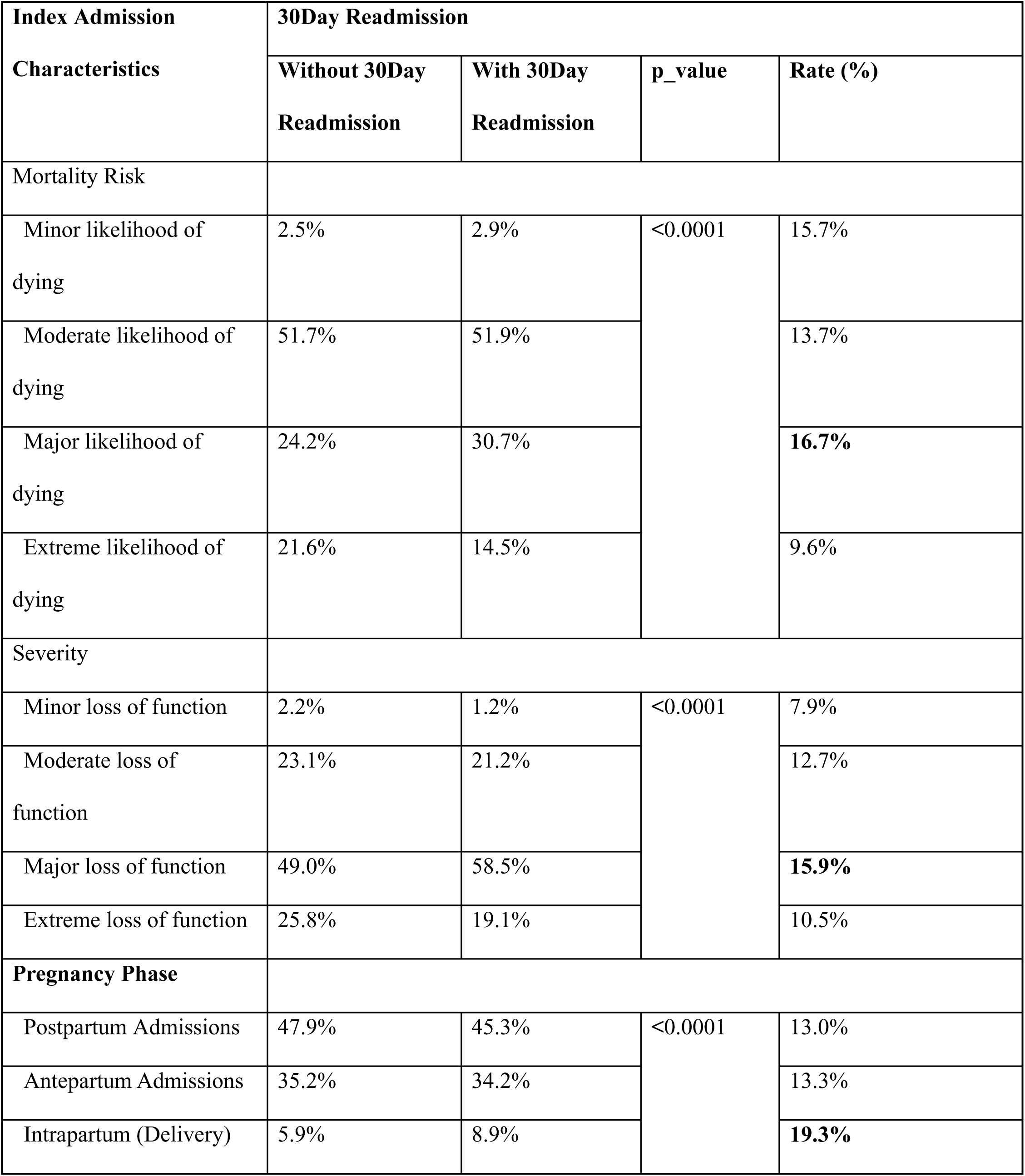
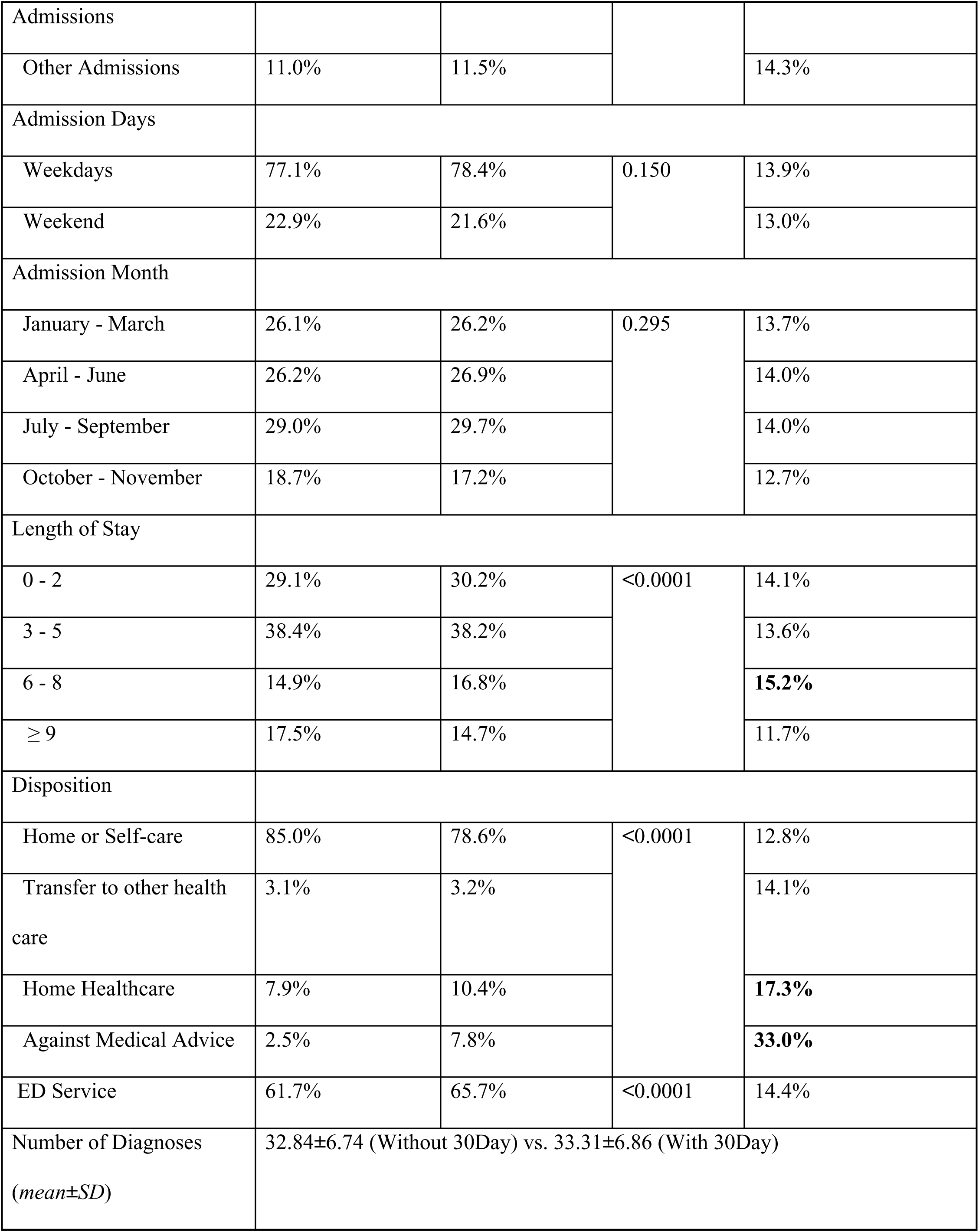
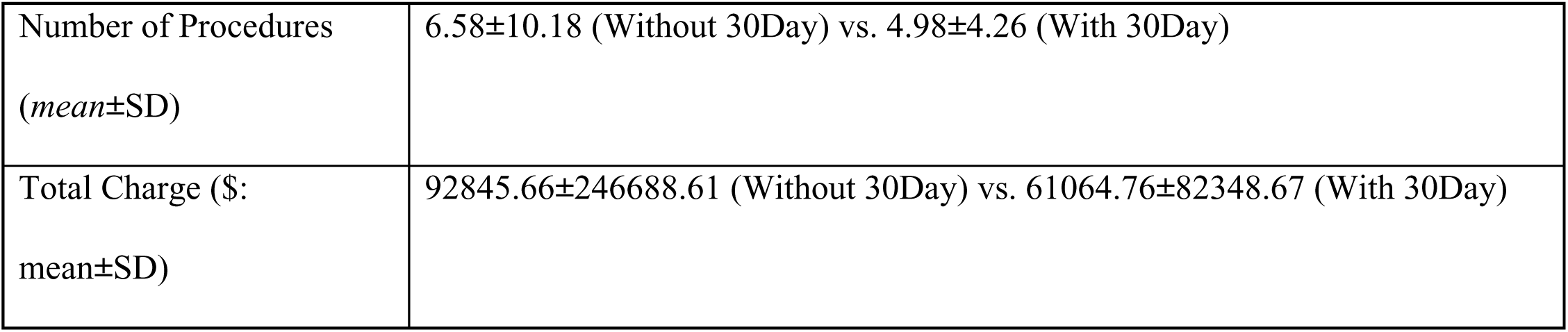
Index Admission Characteristics, Hospitalization Patterns, and Discharge Disposition by 30-Day Readmission Status.

| Index Admission | 30Day Readmission |  |  |  |
| --- | --- | --- | --- | --- |
| Characteristics | Without 30Day<br>Readmission | With 30Day<br>Readmission | p_value | Rate (%) |
| Mortality Risk |  |  |  |  |
| Minor likelihood of<br>dying | 2.5% | 2.9% | <0.0001 | 15.7% |
| Moderate likelihood of<br>dying | 51.7% | 51.9% |  | 13.7% |
| Major likelihood of<br>dying | 24.2% | 30.7% |  | **16.7%** |
| Extreme likelihood of<br>dying | 21.6% | 14.5% |  | 9.6% |
| Severity |  |  |  |  |
| Minor loss of function | 2.2% | 1.2% | <0.0001 | 7.9% |
| Moderate loss of<br>function | 23.1% | 21.2% |  | 12.7% |
| Major loss of function | 49.0% | 58.5% |  | **15.9%** |
| Extreme loss of function | 25.8% | 19.1% |  | 10.5% |
| Pregnancy Phase |  |  |  |  |
| Postpartum Admissions | 47.9% | 45.3% | <0.0001 | 13.0% |
| Antepartum Admissions | 35.2% | 34.2% |  | 13.3% |
| Intrapartum (Delivery) | 5.9% | 8.9% |  | **19.3%** |
| Admissions |  |  |  |  |
| Other Admissions | 11.0% | 11.5% |  | 14.3% |
| Admission Days |  |  |  |  |
| Weekdays | 77.1% | 78.4% | 0.150 | 13.9% |
| Weekend | 22.9% | 21.6% |  | 13.0% |
| Admission Month |  |  |  |  |
| January - March | 26.1% | 26.2% | 0.295 | 13.7% |
| April - June | 26.2% | 26.9% |  | 14.0% |
| July - September | 29.0% | 29.7% |  | 14.0% |
| October - November | 18.7% | 17.2% |  | 12.7% |
| Length of Stay |  |  |  |  |
| 0 - 2 | 29.1% | 30.2% | <0.0001 | 14.1% |
| 3 - 5 | 38.4% | 38.2% |  | 13.6% |
| 6 - 8 | 14.9% | 16.8% |  | **15.2%** |
| ≥ 9 | 17.5% | 14.7% |  | 11.7% |
| Disposition |  |  |  |  |
| Home or Self-care | 85.0% | 78.6% | <0.0001 | 12.8% |
| Transfer to other health care | 3.1% | 3.2% |  | 14.1% |
| Home Healthcare | 7.9% | 10.4% |  | **17.3%** |
| Against Medical Advice | 2.5% | 7.8% |  | **33.0%** |
| ED Service | 61.7% | 65.7% | <0.0001 | 14.4% |
| Number of Diagnoses<br>(*mean±SD*) | 32.84±6.74 (Without 30Day) vs. 33.31±6.86 (With 30Day) |  |  |  |

|  |  |
| --- | --- |
| Number of Procedures<br>(*mean*±SD) | 6.58±10.18 (Without 30Day) vs. 4.98±4.26 (With 30Day) |
| Total Charge (\$:<br>mean±SD) | 92845.66±246688.61 (Without 30Day) vs. 61064.76±82348.67 (With 30Day) |

### Comorbidities

As seen in Table 3, comorbid diagnoses as assessed by the Elixhauser Comorbidity Score including renal failure, solid tumors, peptic ulcer disease with bleeding, and diabetes with complications carried the highest readmission rates. Pregnancy-related comorbidities were associated with significant increased rate of readmission, with substance use or addiction (17.1%, *p= <0.0001*), psychosocial circumstances (23.7%, p <0.0001) and behavioral conditions (21.8%, p= *< 0.0001*) carrying the highest risk.

**Table 3.**
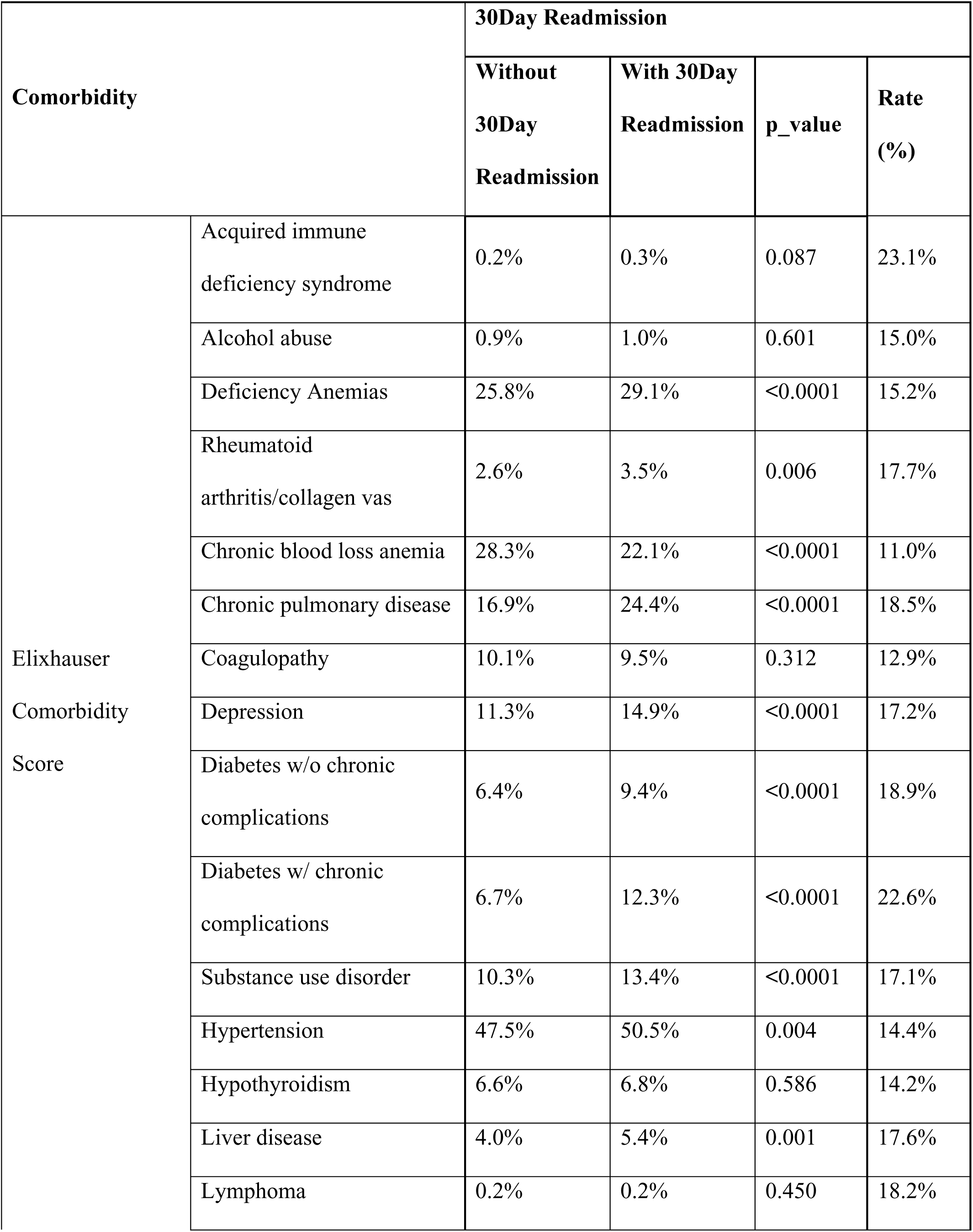
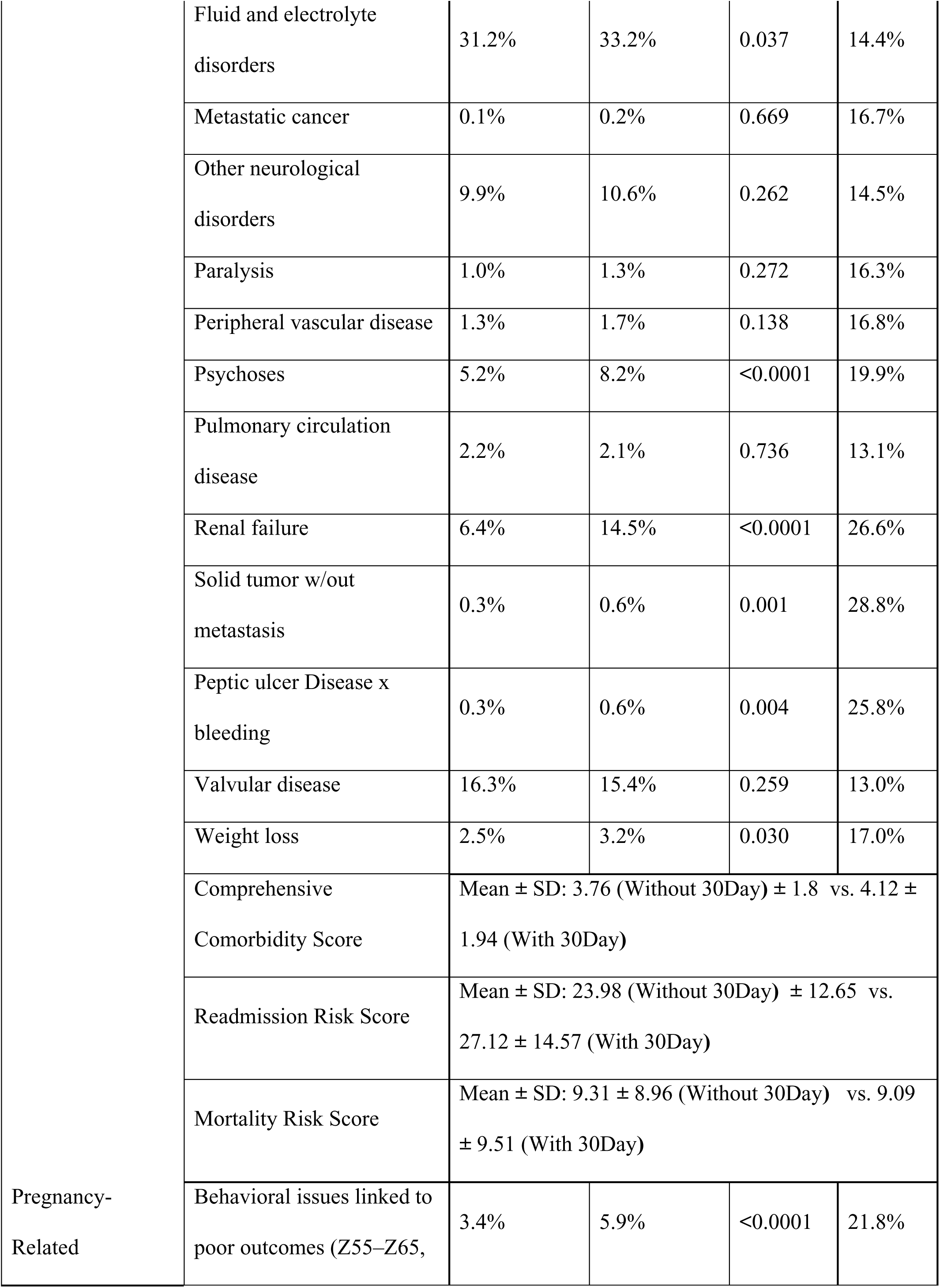
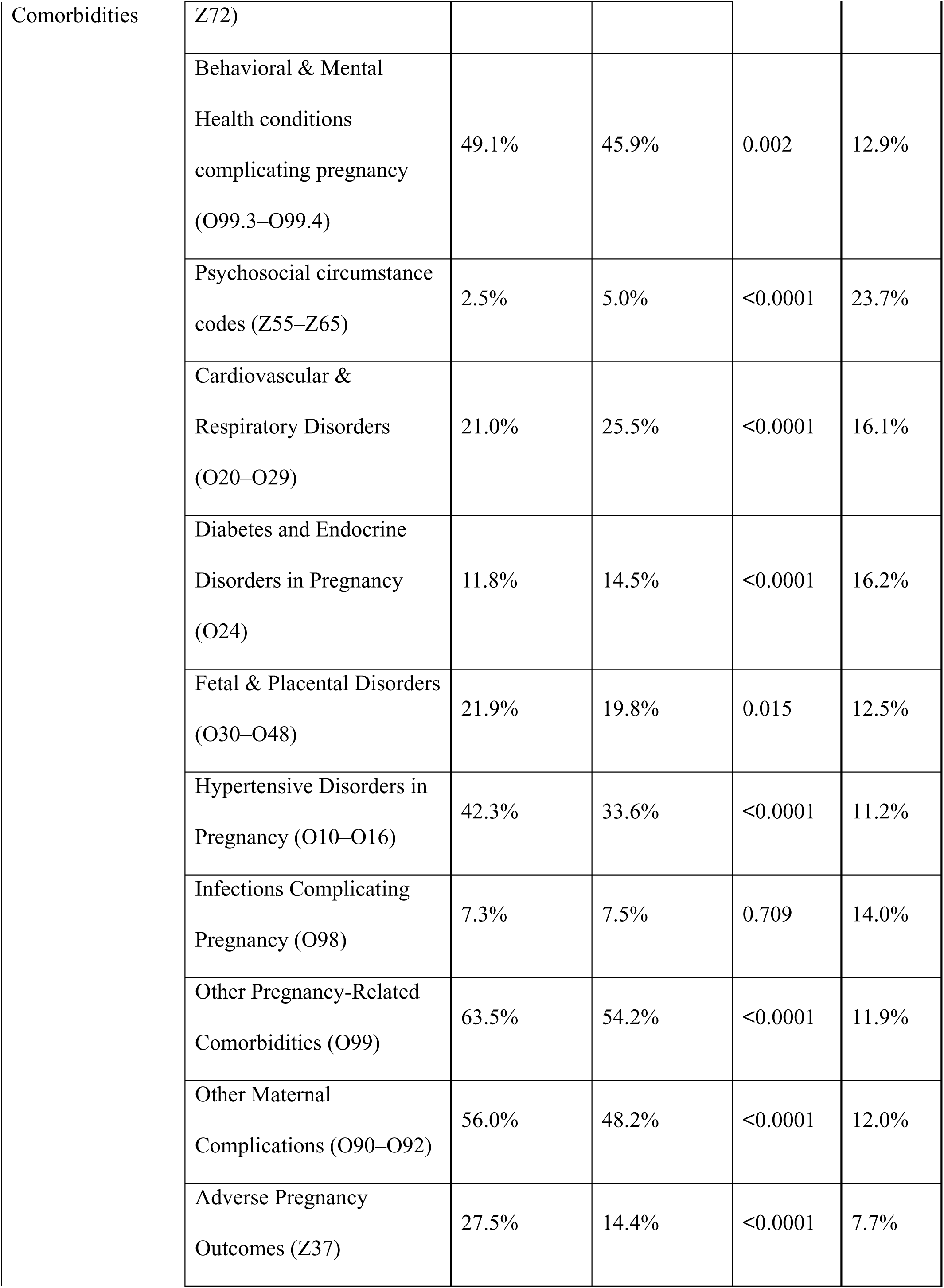
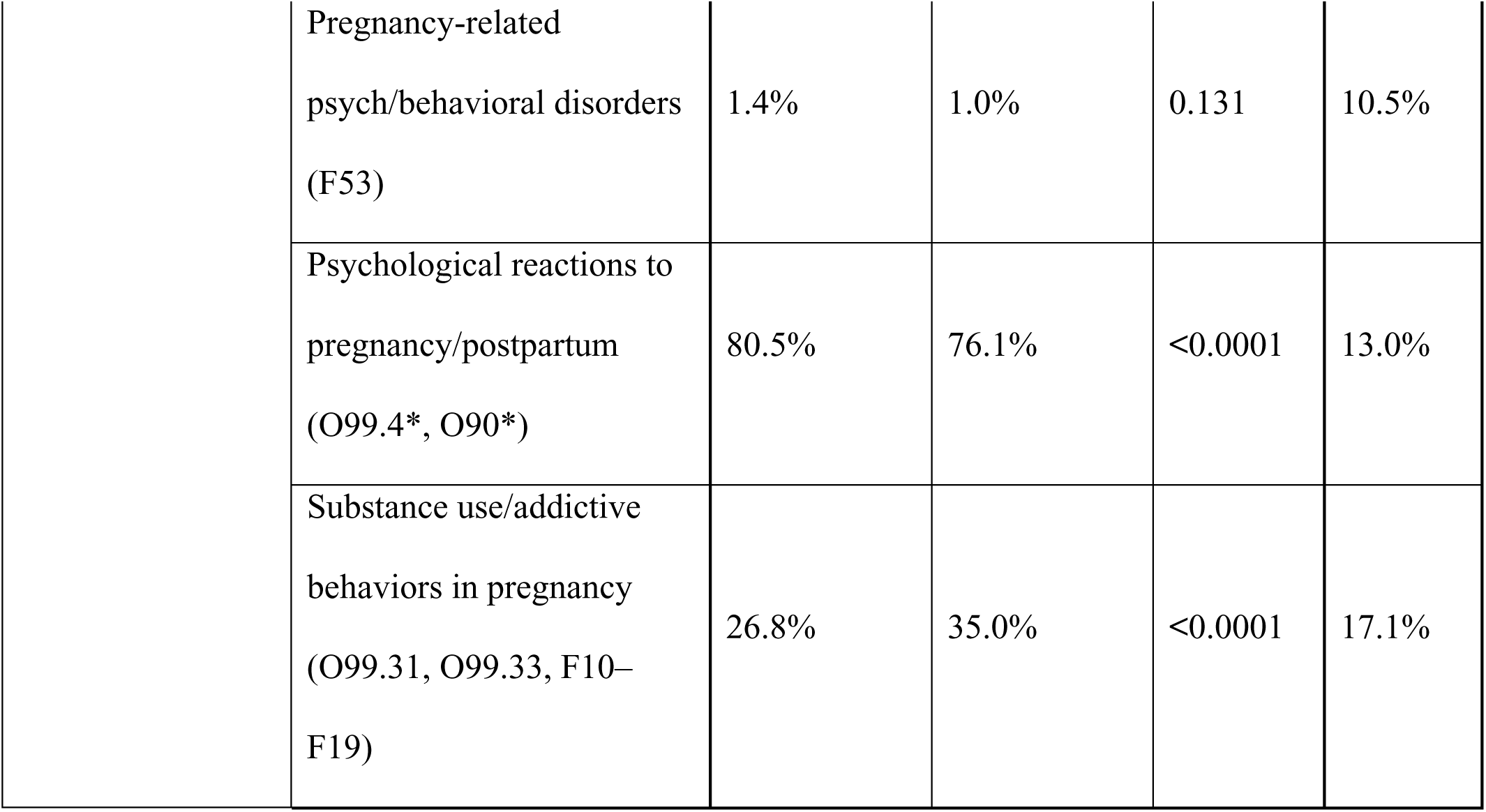
Elixhauser Comorbidities and Pregnancy-Related Comorbidities Associated with 30-Day Readmission, Including Prevalence, Readmission Rates, and Statistical Significance.

### Major Diagnostic Categories of Index Admissions

Figures 2 and 3 represent the distribution of Major Diagnostic Categories (MDCs) for index admissions, stratified by whether a 30-day readmission occurred. Our cohort includes women aged 13–49 years with an index hospitalization that was pregnancy-related (antepartum, intrapartum, or postpartum) and in which HF was coded as either a primary or secondary diagnosis. While pregnancy-related status and the presence of HF define cohort eligibility, MDCs reflect the principal reason for hospitalization, which may vary across patients. The Major Diagnostic Categories (MDC) for index admission among patients with a 30-day readmission are shown in Figure 2. Pregnancy, childbirth, and peripartum accounted for nearly 60% of index admissions leading to readmission. Circulatory and mental health diagnoses comprised much of the remainder (20%). Of the MDCs attributed to the circulatory system, the majority were coded as heart failure and shock (56.6%), followed by ischemic / acute coronary syndrome (14%).

**Figure 2.**
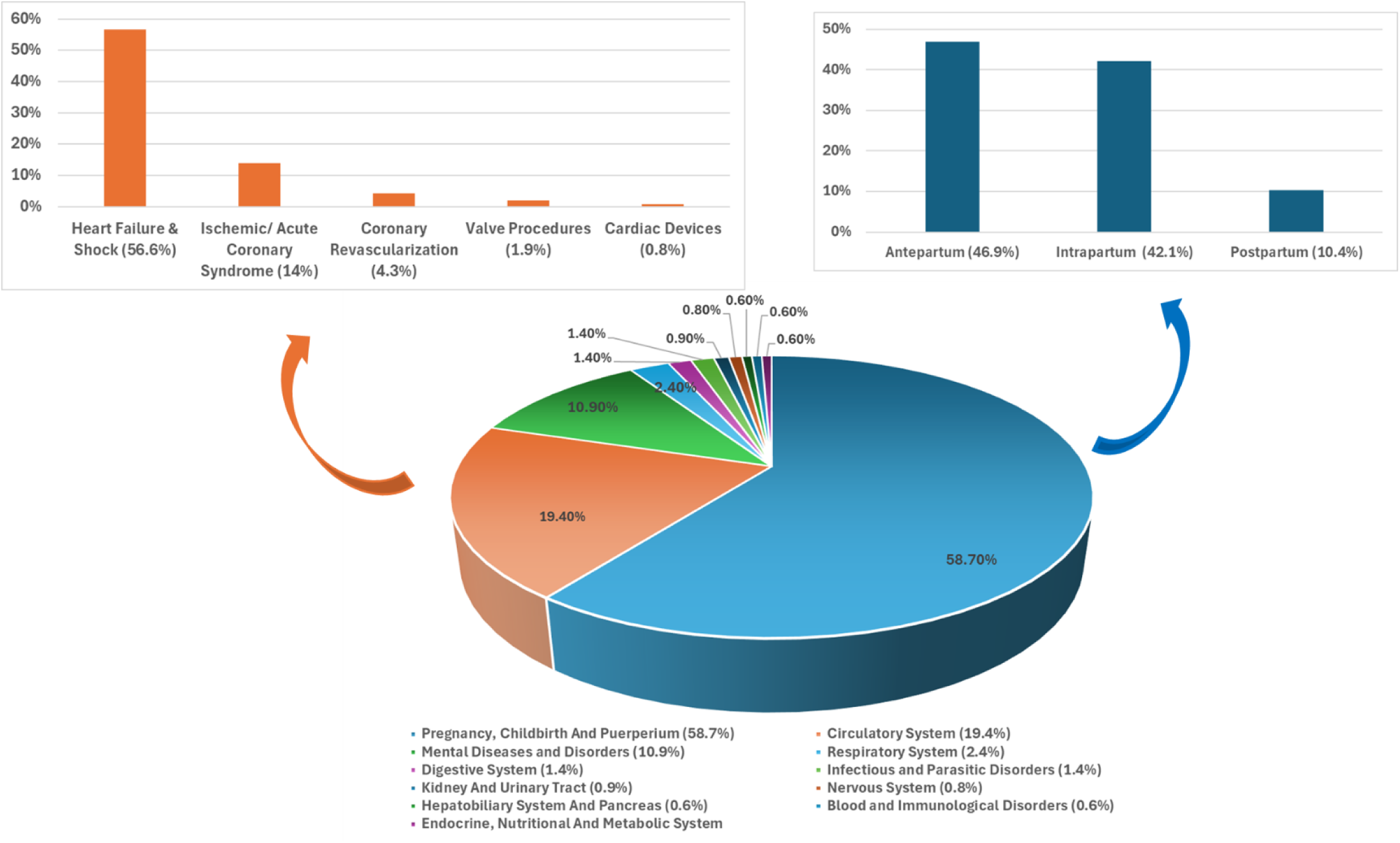
Distribution of Major Diagnostic Categories (MDCs) for Index Admissions Among Patients With 30-Day Readmission.

**Figure 3.**
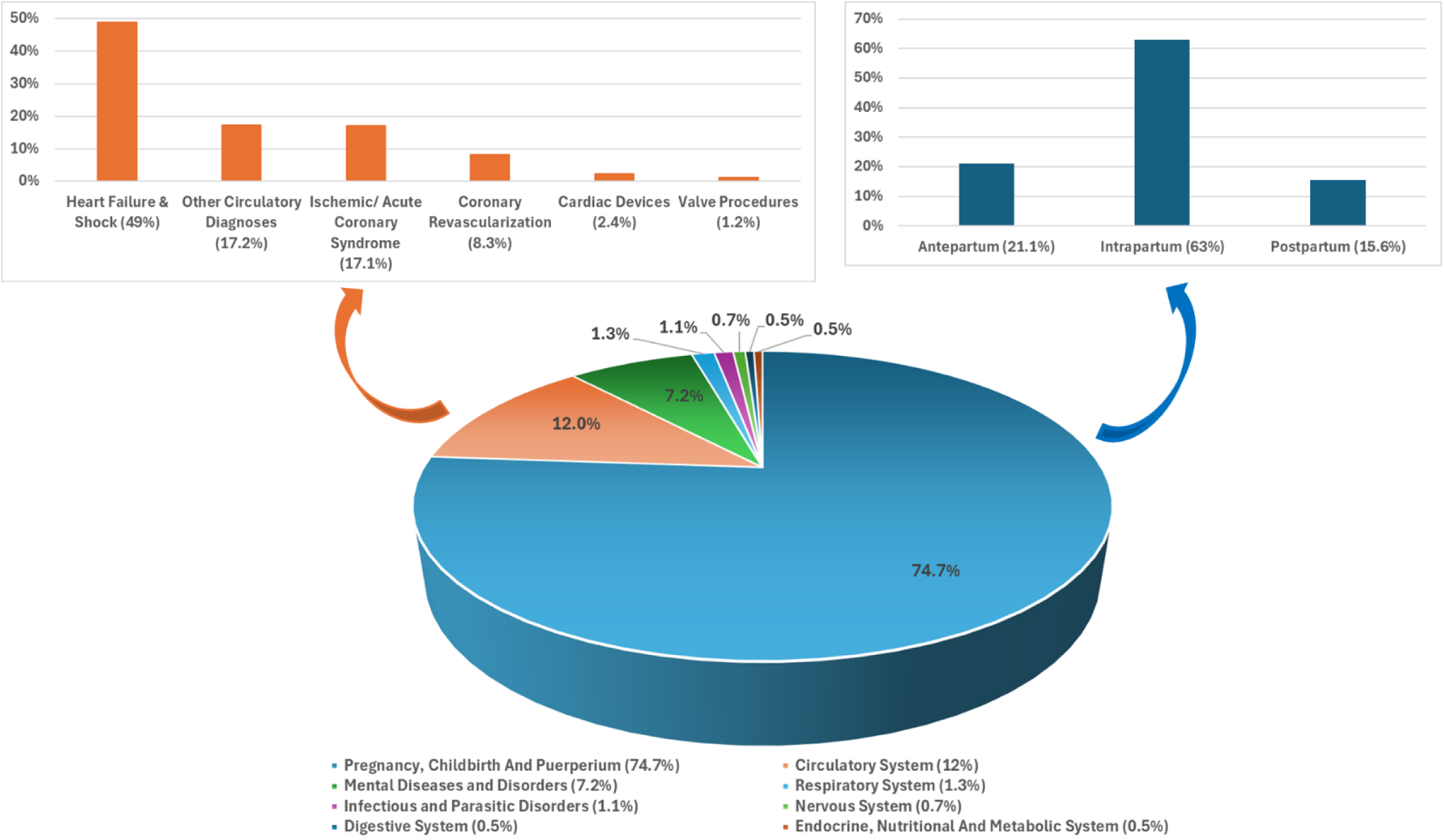
Distribution of Major Diagnostic Categories (MDCs) for Index Admissions Among Patients Without 30-Day Readmission.

In contrast, in those without a 30-day readmission, a higher proportion (75%) were admitted for pregnancy-related diagnoses, and circulatory causes represented only 12% of MDCs (Figure 3).

Figure 4 displays the MDCs for the 30-day readmissions. Approximately 50% of MDCs were related to pregnancy, most of these specifically due to delivery (54.2%). Diseases of the circulatory system made up most of the remainder of MDCs, at 26.19%. Heart failure and shock accounted for more than half of the MDC for readmission attributed to a circulatory system.

**Figure 4.**
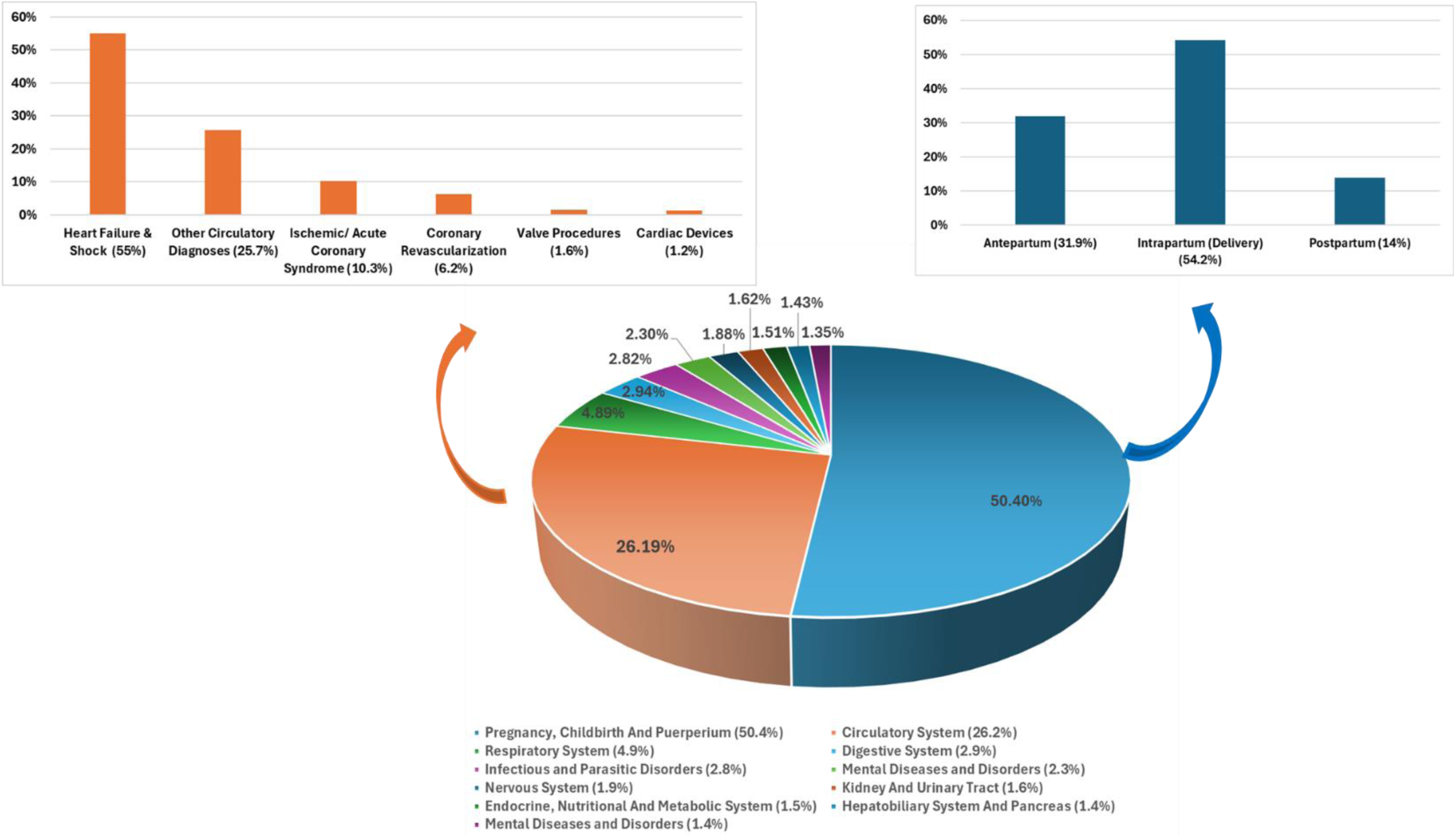
Etiology of 30-Day Readmissions, Including Circulatory and Pregnancy-Related DRGs.

### Timing of Readmissions

Most readmission occurred between days 6–8, and more than half of all readmissions occurred within the first 16 days post-discharge (Figure 5).

**Figure 5.**
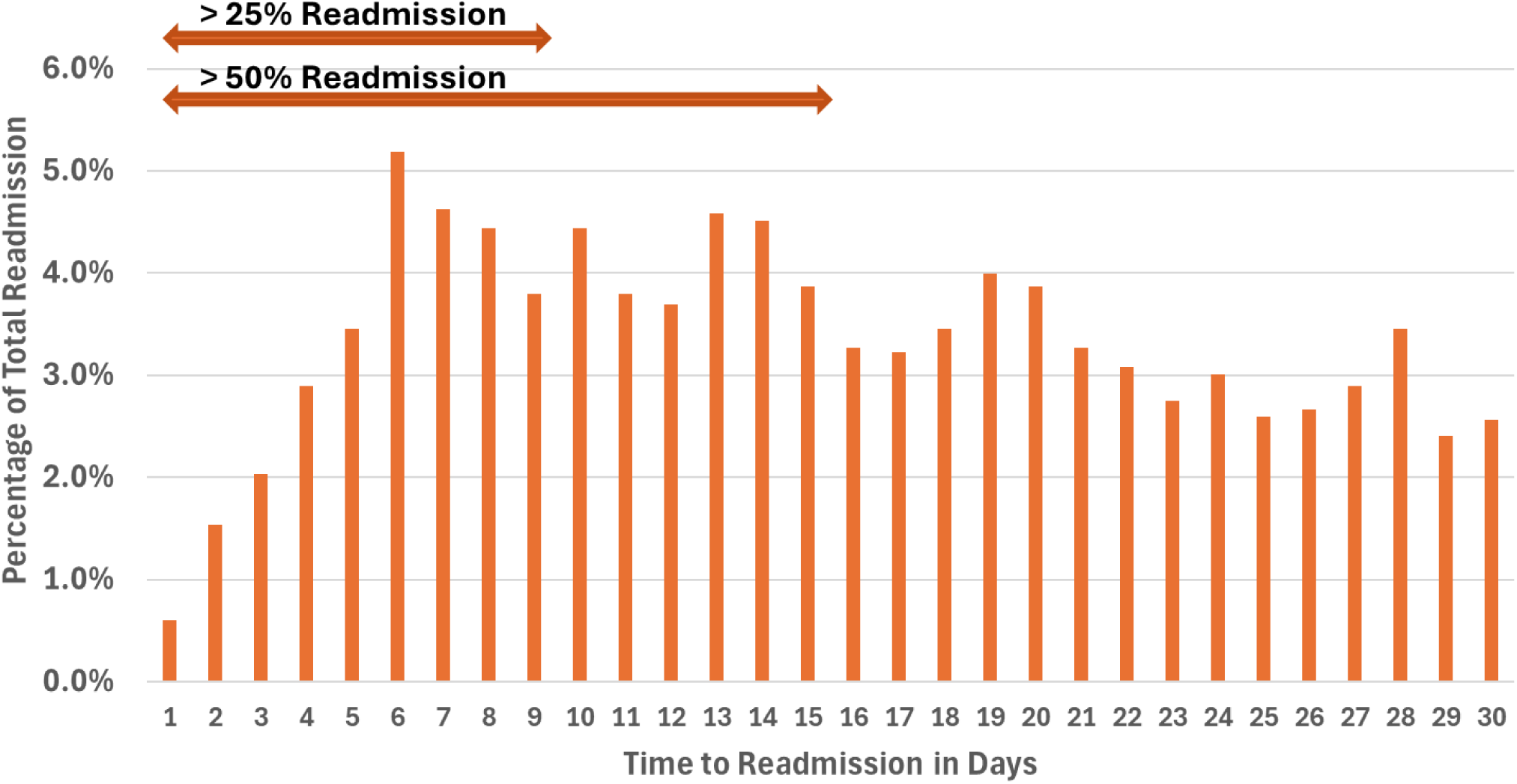
Timing of 30-Day Readmission After Discharge: Percentage of Total Readmissions by Day.

## DISCUSSION

The purpose of this study was to investigate HF readmissions as they relate to pregnancy in a modern cohort of reproductive-aged women. In this study, we found that HF readmission rates in pregnancy are high and involve a spectrum of HF subtypes. The highest risk of readmission occurred after an index hospitalization at the time of delivery and re-admission risk increased with increasing Length of Stay (LOS) and maternal age. Socioeconomic factors and behavioral patterns including substance use portend much higher readmission rates.

HF readmissions have significant economic consequences and lead to worse health outcomes.^19–21^ The overall readmission rate following pregnancy-related HF admissions in our study was 13.7%. PPCM has largely been the prototypical condition for the study of HF in the pregnant population, with varying reports of effect on readmission, and a focus on systolic HF. One study that looked at 90-day readmission trends found that among women discharged after a delivery complicated by PPCM, approximately 0.05% were readmitted with a new or worsening diagnosis of PPCM.^22^ Another 20-year population study found that 75% of women with PPCM were re-hospitalized for any cause at least once.^23^ Additional studies reported 30-day readmission rates ranging from 13-15%.^24,25^ In this study, we found that systolic heart failure and combined heart failure were most likely to result in a readmission at 30 days with readmission rates of 14.8 and 16.1%, respectively, compared to diastolic or other HF. Patients with systolic HF had no difference when coded as PPCM, and when PPCM was examined separately, there was no significant difference in readmission rates. These findings highlight the importance of studying different types of HF phenotypes that may affect pregnancy outside of PPCM, including preexisting HF states along the pregnancy continuum.^10,26,27^

Index admissions for delivery were most likely to be followed by a 30-day readmission compared to other phases of pregnancy. This indicates that this period was the most acutely destabilizing time for HF patients in our cohort and suggests the need to consider earlier and more frequent follow up post-discharge to mitigate this pattern. Readmissions following hospitalization for pregnancy-related HF also increased with increasing maternal age. This helps further characterize high-risk cohorts that require specialized care. The CDC released data confirming that while birth rates for younger women are falling, rates for those aged 40-44 are increasing.^28^ Another report found that the percentage of first births among mothers aged 35 and older increased by 25% between 2016 and 2023.^29^ With a growing number of women having children at an older age, there is an increasing number of patients in this high-risk cohort with higher complications and risk of hospital readmission. Our data indicates increasing readmissions in patients with pregnancy related HF in older age groups and furthers the argument for targeted interventions and research in this vulnerable population.

A longer LOS, specifically 6-8 days, conferred the highest readmission rates in our cohort at 15.7%. This is in contrast with the general HF literature, where LOS is often inversely related to readmission rates. The presumption in the general HF population is that the push toward decreasing length of stay to reduce hospital cost ultimately results in patients being discharged before they are fully diuresed or have achieved euvolemia, leading to increased likelihood that they will be readmitted. Further studies are needed to investigate why in patients with pregnancy-related HF, the relationship between LOS and readmission rates differs from that seen in the general HF population.^30,31^

Other factors that resulted in higher readmission rates were lower socioeconomic status (SES) and alcohol or substance use disorder. The readmission rates for substance use disorder and alcohol use were 17.2% and 18.6%, respectively. Similarly, patients that were discharged against medical advice had the highest readmission rate at 33%. This suggests that social determinants of health are stronger predictors than clinical factors alone. These risk factors have already been well-established in general HF patients. A 2025 scoping review showed that patients with low SES experience 24% higher 30-day readmission rates due to financial instability and restricted access to care. Low health literacy was also found to increase readmission risk by 27%.^32^ Another recent study demonstrated that substance use was present in 38% of patients with congestive heart failure admitted at a safety net hospital and was associated with higher 30-day readmissions.^33^ Various interventions have attempted to address social determinants of health in the pregnant population specifically. This has been one driver of the development of cardio-obstetric programs, with one goal being to address social determinants of health. Other initiatives have included the American College of Gynecology (ACOG) Safe Motherhood initiative, Alliance for Innovation on Maternal Health (AIM), and “See you in 7”.^34–36^ There is no data on these types of programs and interventions on readmissions for pregnancy-related HF.

The timing of readmission in our study was highest within 1-2 weeks, with more than half of all readmissions occurring within the first 16 days post discharge. This pattern is similar to findings in a general HF population admitted with either a primary or secondary diagnosis of HF.^37^ Because of the high economic burden of general HF readmissions specifically,^38,39^ decreasing the rate of readmissions in this population has long been a policy priority. The ‘within 7 days’ timing for post discharge follow up in general HF patients was first established as a national target in 2009 by the Hospital to Home initiative mentioned above.^36^ This initiative was further supported by clinical data that looked at the associations between outpatient follow up within 7 days after discharge and readmission within 30 days. Patients with higher early follow up rates were found to have a lower risk of 30-day readmission.^40^ Thus the 7-day follow up window has been established as the standard of care. Our data suggests that post discharge follow up within 7 days should be extended to patients with a diagnosis of pregnancy-related HF, with consideration to extend a high level of vigilance to 2 weeks after discharge based on the readmission patterns reported in this study.

HF and shock were the highest MDCs outside of pregnancy attributable causes, accounting for 56.6% of MDCs related to the circulatory system. Cardiogenic shock (CS) is defined as a complex syndrome characterized by inadequate tissue perfusion due to reduced cardiac output resulting from a wide array of underlying causes.^41,42^ The mortality rate in CS remains high at 40-50% despite several advances in cardiovascular critical care.^43–45^ The investigation of this high risk cohort is limited by the administrative nature of this database. Further studies are necessary to investigate the severity of HF and the percentage of readmissions made up of shock, specifically in patients along the pregnancy continuum, representing a population at high risk for morbidity and mortality.

There are limitations of this study inherent to the administrative nature of the database used. Coding errors may affect the accurate classification of HF diagnoses or pregnancy-related comorbidities. The study also lacks clinical data such as echocardiography, medications, and outpatient clinical visits which would likely aid in the interpretation of 30-day readmission rates. Further studies are warranted to better define this complex and high-risk population to allow for development of optimal care pathways. Despite the limitations, the NRD administrative database has been validated in population health studies and has a large sample representing all payers in the United States.

## CONCLUSIONS

This study characterizes the 30-day readmission patterns after admission complicated by pregnancy-related HF. The overall readmission rate of our cohort was 13.7% and significantly higher among specific subgroups. Predictors are largely driven by a combination of clinical complexity, timing of HF relative to delivery and social vulnerabilities, revealing the need for a better understanding of how to provide patient-specific multidisciplinary transitional care, especially within the critical 1-2 weeks post discharge.

## Data Availability

The National Readmission Database was used

https://hcup-us.ahrq.gov/nrdoverview.jsp

## ACKNOWLEDGEMENTS

None

## SUPPLEMENTAL MATERIAL

Supplemental Table1. Cohort Definition Diagnosis Codes (ICD-10-CM and DRG Codes)

Supplemental Table2. Major Diagnostic Categories (MDCs)

Supplemental Table3. MDC-Associated DRGs Used in Etiology Analyses

Supplemental Table4. Pregnancy-Related Comorbidities (ICD-10-CM)

Supplemental Table5. Behavioral or Social Factors Influencing Pregnancy Outcomes (ICD-10-CM)

## REFERENCES

1. Williamson CG, Altendahl M, Martinez G, et al. Cardiovascular Disease in Pregnancy. JACC: Advances. 2024;3(8):101071. doi:10.1016/j.jacadv.2024.101071

2. Quiñones JN, Walheim L, Mann K, Rochon M, Ahnert AM. Impact of type of maternal cardiovascular disease on pregnancy outcomes among women managed in a multidisciplinary cardio-obstetrics program. American Journal of Obstetrics & Gynecology MFM. 2021;3(4):100377. doi:10.1016/j.ajogmf.2021.100377

3. Ramlakhan KP, Johnson MR, Roos-Hesselink JW. Pregnancy and cardiovascular disease. Nat Rev Cardiol. 2020;17(11):718–731. doi:10.1038/s41569-020-0390-z

4. Tenner S, Vege SS, Sheth SG, et al. American College of Gastroenterology Guidelines: Management of Acute Pancreatitis. Am J Gastroenterol. 2024;119(3):419–437. doi:10.14309/ajg.0000000000002645

5. CDC. Centers for Disease Control and Prevention (CDC). “Pregnancy-Related Deaths: Data from Maternal Mortality Review Committees.” Published online August 22, 2025. https://www.cdc.gov/maternal-mortality/php/data-research/mmrc/index.html

6. Mehta LS, Warnes CA, Bradley E, et al. Cardiovascular Considerations in Caring for Pregnant Patients: A Scientific Statement From the American Heart Association. Circulation. 2020;141(23). doi:10.1161/CIR.0000000000000772

7. Lau ES, D’Souza V, Zhao Y, et al. Contemporary Burden of Cardiovascular Disease in Pregnancy: Insights From a Real-World Pregnancy Electronic Health Record Cohort. Circulation. 2025;152(15):1044–1055. doi:10.1161/CIRCULATIONAHA.125.074692

8. Declercq E, Zephyrin LC. Maternal Mortality in the United States, 2025. Published online 2025. doi:10.26099/KDFD-FC19

9. Centers for Disease Control and Prevention (CDC). Pregnancy Mortality Surveillance System (PMSS). Maternal Mortality Prevention. Updated 2024. Available at: https://www.cdc.gov/maternal-mortality/php/surveillance/index.html

10. Mogos MF, Piano MR, McFarlin BL, Salemi JL, Liese KL, Briller JE. Heart Failure in Pregnant Women: A Concern Across the Pregnancy Continuum. Circ: Heart Failure. 2018;11(1):e004005. doi:10.1161/CIRCHEARTFAILURE.117.004005

11. Ruys TPE, Roos-Hesselink JW, Hall R, et al. Heart failure in pregnant women with cardiac disease: data from the ROPAC. Heart. 2014;100(3):231–238. doi:10.1136/heartjnl-2013-304888

12. Pfaller B, Sathananthan G, Grewal J, et al. Preventing Complications in Pregnant Women With Cardiac Disease. Journal of the American College of Cardiology. 2020;75(12):1443–1452. doi:10.1016/j.jacc.2020.01.039

13. Roos-Hesselink J, Baris L, Johnson M, et al. Pregnancy outcomes in women with cardiovascular disease: evolving trends over 10 years in the ESC Registry Of Pregnancy And Cardiac disease (ROPAC). European Heart Journal. 2019;40(47):3848–3855. doi:10.1093/eurheartj/ehz136

14. Girsen AI, Leonard SA, Butwick AJ, Joudi N, Carmichael SL, Gibbs RS. Early postpartum readmissions: identifying risk factors at birth hospitalization. AJOG Global Reports. 2022;2(4):100094. doi:10.1016/j.xagr.2022.100094

15. Rosenfeld EB, Brandt JS, Fields JC, et al. Chronic Hypertension and the Risk of Readmission for Postpartum Cardiovascular Complications. Obstet Gynecol. 2023;142(6):1431–1439. doi:10.1097/AOG.0000000000005424

16. Nizamuddin J, Gupta A, Patel V, et al. Hypertensive Diseases of Pregnancy Increase Risk of Readmission With Heart Failure: A National Readmissions Database Study. Mayo Clinic Proceedings. 2019;94(5):811–819. doi:10.1016/j.mayocp.2018.08.032

17. Jarvie JL, Metz TD, Davis MB, Ehrig JC, Kao DP. Short-term risk of cardiovascular readmission following a hypertensive disorder of pregnancy. Heart. 2018;104(14):1187–1194. doi:10.1136/heartjnl-2017-312299

18. HCUP. Agency for Healthcare Research and Quality, Healthcare Cost and Utilization Project. Elixhauser comorbidity software refined for ICD-10-CM. Accessed September 21, 2021. https://www.hcup-us.ahrq.gov/toolssoftware/comorbidityicd10/comorbidity_icd10.jsp

19. Kum Ghabowen I, Epane JP, Shen JJ, Goodman X, Ramamonjiarivelo Z, Zengul FD. Systematic Review and Meta-Analysis of the Financial Impact of 30-Day Readmissions for Selected Medical Conditions: A Focus on Hospital Quality Performance. Healthcare. 2024;12(7):750. doi:10.3390/healthcare12070750

20. Beauvais B, Whitaker Z, Kim F, Anderson B. Is the Hospital Value-Based Purchasing Program Associated with Reduced Hospital Readmissions? JMDH. 2022;Volume 15:1089–1099. doi:10.2147/JMDH.S358733

21. Foroutan F, Rayner DG, Ross HJ, et al. Global Comparison of Readmission Rates for Patients With Heart Failure. Journal of the American College of Cardiology. 2023;82(5):430–444. doi:10.1016/j.jacc.2023.05.040

22. Masoomi R, Shah Z, Dawn B, Gupta K. P6190Incidence of peripartum cardiomyopathy diagnosed in 90-day post-delivery: insights from the nationwide readmission database. European Heart Journal. 2017;38(suppl_1). doi:10.1093/eurheartj/ehx493.P6190

23. Jackson AM, Macartney M, Brooksbank K, et al. A 20-year population study of peripartum cardiomyopathy. European Heart Journal. 2023;44(48):5128–5141. doi:10.1093/eurheartj/ehad626

24. Shah M, Ram P, Lo KB, et al. Etiologies, Predictors, and Economic Impact of 30-Day Readmissions Among Patients With Peripartum Cardiomyopathy. The American Journal of Cardiology. 2018;122(1):156–165. doi:10.1016/j.amjcard.2018.03.018

25. Chhabra N, Gupta A, Chibber R, et al. Outcomes and mortality in parturient and non-parturient patients with peripartum cardiomyopathy: A national readmission database study. Pregnancy Hypertension. 2017;10:143–148. doi:10.1016/j.preghy.2017.07.147

26. Ersilia M. DeFilippis, Catriona Bhagra, Jillian Casale, Patricia Ging, Francesca Macera, Lynn Punnoose, Kismet Rasmusson, Garima Sharma, Karen Sliwa, Sara Thorne, Mary Norine Walsh, and Michelle M. Kittleson. Cardio-Obstetrics and Heart Failure: JACC: Heart Failure State-of-the-Art Review. JACC: Heart Failure *Volume* 11, *Number 9 5,431*. Published online September 5, 2023.

27. Lee SU, Park JY, Hong S, et al. Risk factors for pregnancy-associated heart failure with preserved ejection fraction and adverse pregnancy outcomes: a cross-sectional study. BMC Pregnancy Childbirth. 2024;24(1):211. doi:10.1186/s12884-024-06402-5

28. Joyce A. Martin, M.P.H., Brady E. Hamilton, Ph.D., and Michelle J.K. Osterman, M.H.S. CDC National Center for Health Statistics. (2025). Number 535: Births in the United States, 2024.

29. Andrea D. Brown, Ph.D., M.P.H; Brady E. Hamilton, Ph.D.; Dmitry M. Kissin, M.D., M.P.H.; and, Joyce A. Martin, M.P.H. Trends in Mean Age of Mothers: United States, 2016–2023.“ National Vital Statistics Reports, Vol. 74, No. 9. Published June 13, 2025. https://www.cdc.gov/nchs/data/nvsr/nvsr74/nvsr74-09.pdf

30. Khan H, Greene SJ, Fonarow GC, et al. Length of Hospital Stay and 30-Day Readmission Following Heart Failure Hospitalization: Insights from the EVEREST Trial. European Journal of Heart Failure. 2015;17(10):1022–1031. doi:10.1002/ejhf.282

31. Sud M, Yu B, Wijeysundera HC, et al. Associations Between Short or Long Length of Stay and 30-Day Readmission and Mortality in Hospitalized Patients With Heart Failure. JACC: Heart Failure. 2017;5(8):578–588. doi:10.1016/j.jchf.2017.03.012

32. Duran K, Copel LC, Hinkle JL. Examining the implications of social determinants of health on hospital readmission for patients with heart failure: a scoping review. Discov Soc Sci Health. 2025;5(1):121. doi:10.1007/s44155-025-00284-4

33. Majdalani R, AlShammari A, Thearle M, et al. Prevalence and Impact of Substance Use on Hospitalization and Post-Discharge Outcomes in Individuals with Congestive Heart Failure: Findings from a Safety-Net Hospital. IJERPH. 2025;22(12):1832. doi:10.3390/ijerph22121832

34. Care for Pregnant and Postpartum People with Substance Use Disorder. (2024 Update). Council on Patient Safety in Women’s Health Care. https://saferbirth.org/patient-safety-bundles/

35. American College of Obstetricians and Gynecologists District II. Safe Motherhood Initiative: Hypertensive Disorders of Pregnancy. https://www.acog.org/community/districts-and-sections/district-ii/programs-and-resources/safe-motherhood-initiative

36. American College of Cardiology. Hospital to Home (H2H) “See You in 7” Challenge. https://cvquality.acc.org/initiatives/hospital-to-home/Projects/see-you-in-7

37. Kim MJ, Aseltine RH, Tabtabai SR. Understanding the Burden of 30-Day Readmission in Patients With Both Primary and Secondary Diagnoses of Heart Failure: Causes, Timing, and Impact of Co-Morbidities. The American Journal of Cardiology. 2024;210:76–84. doi:10.1016/j.amjcard.2023.09.086

38. Kwok CS, Abramov D, Parwani P, et al. Cost of inpatient heart failure care and 30-day readmissions in the United States. International Journal of Cardiology. 2021;329:115–122. doi:10.1016/j.ijcard.2020.12.020

39. Urbich M, Globe G, Pantiri K, et al. A Systematic Review of Medical Costs Associated with Heart Failure in the USA (2014–2020). PharmacoEconomics. 2020;38(11):1219–1236. doi:10.1007/s40273-020-00952-0

40. Hernandez AF. Relationship Between Early Physician Follow-up and 30-Day Readmission Among Medicare Beneficiaries Hospitalized for Heart Failure. JAMA. 2010;303(17):1716. doi:10.1001/jama.2010.533

41. Lüsebrink E, Binzenhöfer L, Adamo M, et al. Cardiogenic shock. The Lancet. 2024;404(10466):2006–2020. doi:10.1016/S0140-6736(24)01818-X

42. Waksman R, Pahuja M, Van Diepen S, et al. Standardized Definitions for Cardiogenic Shock Research and Mechanical Circulatory Support Devices: Scientific Expert Panel From the Shock Academic Research Consortium (SHARC). Circulation. 2023;148(14):1113–1126. doi:10.1161/CIRCULATIONAHA.123.064527

43. Karakas M, Akin I, Burdelski C, et al. Single-dose of adrecizumab versus placebo in acute cardiogenic shock (ACCOST-HH): an investigator-initiated, randomised, double-blinded, placebo-controlled, multicentre trial. The Lancet Respiratory Medicine. 2022;10(3):247–254. doi:10.1016/S2213-2600(21)00439-2

44. Ostadal P, Rokyta R, Karasek J, et al. Extracorporeal Membrane Oxygenation in the Therapy of Cardiogenic Shock: Results of the ECMO-CS Randomized Clinical Trial. Circulation. 2023;147(6):454–464. doi:10.1161/CIRCULATIONAHA.122.062949

45. Thiele H, Zeymer U, Akin I, et al. Extracorporeal Life Support in Infarct-Related Cardiogenic Shock. N Engl J Med. 2023;389(14):1286–1297. doi:10.1056/NEJMoa2307227

